# Monitoring fatigue with consumer wearables in multiple sclerosis following rehabilitation: An observational study

**DOI:** 10.1101/2025.11.07.25339665

**Authors:** Andreas M. Baumer, Christina Haag, Chloé Sieber, Marcelo Stegmann, Ramona Sylvester, Roman Gonzenbach, Viktor von Wyl

## Abstract

Fatigue places a great burden on people with multiple sclerosis (PwMS), but fatigue assessments are imperfect and rely on subjective surveys. Assessing digital biomarkers derived from wearable devices has shown promise as an objective measurement method. We investigate their association with fatigue with a special focus on within-person changes and differences between cognitive and motor fatigue. PwMS entering inpatient rehabilitation were recruited and equipped with a consumer-grade fitness watch (Fitbit Inspire HR). Participants were monitored during rehabilitation and four weeks after returning home. We used hierarchical modeling to assess cross-sectional associations between digital biomarkers and fatigue scores and linear models to investigate associations between change scores. In total, 45 PwMS were included in the study (61.7% female, mean age = 46.0 (SD = 8.9). Device wearing compliance was high at 92.0% and participants provided on average 51.6 days of recorded data. The assessed digital biomarkers were generally only weakly and non-significantly associated with fatigue. The strongest cross-sectional association with fatigue was found in intraday variability (b=-0.22, 95%CI = [-0.41, -0.02]), while step counts were most responsive to within-person changes in fatigue (b=-0.32, 95%CI = [-0.68, 0.05]). On average, effect sizes were 23.3% greater for change scores compared to cross-sectional associations and 62.9% greater for motor than for cognitive fatigue. Despite the very consistent data collection by participants, digital biomarkers did not show strong associations with fatigue in MS patients, suggesting general limitations of using consumer-grade wearables for fatigue assessments. We found some evidence that digital biomarkers are better suited to reflect within-person change than interpersonal differences. Additionally, digital biomarkers related more closely to motor than cognitive fatigue. Therefore, the most promising target for future research may be within-person changes in motor fatigue.

## Introduction

Fatigue is the most debilitating and impactful symptom that people with multiple sclerosis (PwMS) face and is experienced by 59.1% of this population (1). Fatigue affects the cognitive and motor functions of PwMS, resulting in forgetfulness, inability to focus or make decisions, as well as muscle soreness, reduced nimbleness, and tiring quickly (2).

Despite this high prevalence and burden, researchers and practitioners still face challenges when assessing and monitoring fatigue. The current gold standard assessment instruments are retrospective self-report surveys, such as the Fatigue Scale for Motor and Cognitive Functions (FSMC; Penner et al., 2009) used in this study. The subjective nature of such self-report surveys introduces challenges for researchers when they try to compare fatigue between patients. Even comparisons within the same patient can be difficult if the survey time points are far apart. To address these challenges, researchers have explored objective and continuous digital biomarkers such as step counts and heart rate using a variety of wearable devices. In previous studies, inertial sensors (4), smart phone-based step counters (5), consumer-grade fitness watches (6,7), and purpose-built actigraphs (8) have all shown some correlations between digital biomarkers and fatigue.

Unfortunately, large interpersonal variability in contextual factors such as physical ability and lifestyle limits how precisely physical activity reflects fatigue symptoms (9). This limitation is more pronounced in PwMS as they exhibit a wide range of disability states, with some PwMS requiring walking aides or wheelchairs. Earlier studies have shown two promising avenues for using digital biomarkers in PwMS. The first approach focuses on digital biomarkers that are independent of overall physical ability, specifically rest-activity-rhythms (RARs). RARs express the strength of a person’s circadian rhythm, and their potential connection to fatigue in multiple sclerosis (MS) was recognized as early as 1993 (10). PwMS have been shown to have disrupted RARs (11) and disruption in RARs has been linked to various deleterious outcomes from cognitive function to mortality (12,13). The second approach to account for baseline differences is to shift to a longitudinal perspective and to focus on intrapersonal changes. Dalla Costa and colleagues as well as Block and colleagues have shown that digital biomarkers can reflect MS progression and may in fact provide a more granular view than traditional clinical measures can (14,15), but evidence remains scarce.

In this study, we investigate how consumer-grade wearables could add to our understanding of fatigue in PwMS, particularly in the later stages of MS progression. We analyzed data from the Barka-MS study, which assessed patient-reported fatigue at three points, at the beginning and end of inpatient rehabilitation as well as four weeks after returning home. As the *first objective*, this study investigates the cross-sectional relationship between digital biomarkers and self-reported fatigue. For our *second objective*, we assess how digital biomarkers respond to changes in fatigue as patients return home after rehabilitation treatment. The *third objective* is to understand whether the relation of digital biomarkers with fatigue differs between cognitive and motor fatigue subscale.

## Methods

### Study setting

The BarKA-MS study took place from January to November 2021 at the Valens Clinics in Switzerland, an in-patient rehabilitation clinic focusing on physical activity, where PwMS typically stay up to four weeks. The aim of the study was to better understand the barriers and enablers for PwMS being physically active, especially after inpatient rehabilitation. Participants were recruited as they entered treatment (week 0) and completed baseline fatigue, and disability assessments. Fatigue assessments were repeated when participants completed rehabilitative treatment (week 4) and four weeks after returning home (week 8). Participants were instructed to wear a Fitbit Inspire HR activity tracker continuously during the eight-week study duration. Sieber and colleagues provide further details on the trial design, and the (Sieber et al., 2023), the study was approved the relevant ethics committee (BASEC# 2020-02350)

### Instruments

#### The fatigue scale for motor and cognitive functions

(FSMC; Penner et al., 2009) is a self-report questionnaire that includes 20 items, which are rated on a 5-point Likert scale. The FSMC consists of a motor fatigue and a cognitive fatigue sub-scale. Sum scores above 42 correspond to mild, those above 52 to moderate, and those above 62 to severe fatigue.

#### The expanded disability status scale

(EDSS; Kurtzke, 1983) is the most commonly used instrument to measure impairment in MS (18). It is the primary scale used to describe disease progression and assess gait challenges in PwMS. It ranges from 0 (no impairment) to 10 (deceased due to MS) in half-point steps, with higher scores indicating higher impairment. We categorized EDSS scores into three groups of ≤3.5, 4-5.5 and ≥6 corresponding to no walking impairment, some walking impairment, and obligatory use of walking aides.

### Digital biomarkers

Fitbit Inspire HR reports step counts, average heart rate estimates in one-minute epochs, and sleep data in 30-second epochs. We obtained and managed the data using the Fitabase service. Polhemus and colleagues investigated the validity of these data in comparison to data gathered by research-grade devices and found that while they were not equivalent, they possessed similar construct validity (19). Because PwMS do not exhibit the movement patterns of healthy adults, fitbit based estimates such as total number of steps, may be less accurate than expected(20).

We focused on commonly used digital biomarkers that have previously been associated with fatigue in PwMS (heart rate, steps, sleep duration, time awake in bed) and the following five RAR metrics. *intraday variability* (IV) measures the fragmentation of circadian rhythm within a day, with lower score showing a lower fragmentation. A range of 0 to 2 has been proposed (21) but is mathematically uncertain (22). *interdaily stability* (IS) indicates stability in activity across multiple days and ranges from 0 to 1, with higher scores indicating greater stability. *relative amplitude* (RA) compares the *ten hours of highest activity* (M10) to the *five hours of lowest activity* (L5) and is a composite measure of physical activity and ability to rest and relax, ranging from 0 to 1. These RARs were calculated on the basis of step counts, further details on the calculations are given by Taphoorn et al. (1993).

Heart rate variability, a commonly referenced biomarker, was unavailable and could not be directly calculated because Fitbit does not report data at such granularity. As a proxy, we exploratively investigated the standard deviation of the provided 1-minute heart rate estimates.

### Statistical analysis

We only considered wearable data if the device was worn for at least ten hours that day to ensure representativeness.

Because the FSMC refers to the past seven days, we derived the biomarkers from wearable data gathered in the week prior to filling out a survey, i.e. the last week of rehabilitation treatment and the fourth week after returning home. This was not possible for the baseline assessment, because patients only started wearing the device at the same time they reported their fatigue. For each patient we therefore obtained biomarkers and fatigue scores at two points in time. For descriptive statistics we calculated means and standard deviations for numeric variables and percentages for categorical variables. For regression analyses, we transformed all variables to z-scores to allow for better comparison.

For *objective 1*, cross-sectional associations between each digital biomarker and fatigue (dependent variable) were calculated, while controlling for age, sex, and EDSS category. We used a hierarchical linear where observations (end of rehabilitation, fourth week of home assessment) were nested within each participant and included a random intercept to account for baseline differences.

For *objective 2*, we constructed a linear model for each digital biomarker. Changes in fatigue score were the dependent variable and changes in the respective digital biomarker an independent variable, alongside age, sex, and EDSS category. We did not use a nested model, as there is only one observation per patient.

For *objective 3*, we expanded the analyses of objectives 1 and 2 with FSMC cognitive and motor subscales as dependent variables.

The analyses were conducted in R using the lme4 (23) package. In general, we emphasized effect size and confidence intervals over statistical significance.

## Results

In total 45 PwMS were recruited into BarKA-MS. For our analysis, one participant was excluded due to faulty Fitbit data. The remaining 44 participants were 61.7% female, with an average age of 46 and different types of MS (Primary-progressive: 8; secondary-progressive: 19; relapsing-remitting: 17). EDSS scores ranged from 2 to 6.5 with a median of 4.0. FSMC total fatigue score assessed when entering rehabilitation treatment was 61.7 (SD=15.4). Demographic information and fatigue metrics of participants at the beginning of treatment are provided in Table 1.

**Table 1.** Demographic and clinical characteristics of participants entering rehabilitation treatment (N = 44)

| Characteristic | N (%) |
| --- | --- |
| Sex female | 29 (61.7%) |
| Multiple Sclerosis Type |  |
| Primary Progressive | 8 (18.2%) |
| Secondary Progressive | 19 (43.2%) |
| Relapsing Remitting | 17 (38.7%) |
| Expanded Disability Status Scale score |  |
| ≤ 3.5 | 12 (27.3%) |
| 4–5.5 | 19 (43.1%) |
| ≥ 6 | 13 (29.5%) |
|  | Mean (SD) |
| Age | 46.0 (8.9) |
| FSMC* when entering rehabilitative treatment |  |
| Motor | 35.4 (6.72) |
| Cognitive | 26.3 (10.9) |
| Total | 61.7 (15.4) |

Note: \*FSMC: Fatigue Scale for Motor and Cognitive function

### Fitbit data quality

On average, the 44 participants had the device for 56.1 study days and wore it for 51.7 days, indicating a compliance of 92.0%. In total, 96.7% of the 2410 days were considered valid with over 10 hours of available data.

### Associations of biometrics with fatigue levels

In total 86 observations from 44 participants (n_clinic_ = 44, n_home_ = 42) were available to investigate the relation of fatigue and biomarkers, as detailed in Table 2. After participants returned home from the clinic, FSMC scores increased by 4.2 points and daily step counts decreased by 2249.8 steps. The RAR biomarkers of IS and RA also increased, indicating the weakening of circadian rhythms.

**Table 2.** Digital biomarkers and fatigue, during the last week of rehabilitation and four weeks after returning home.

| **Fatigue and Digital Biomarkers** | **Clinic – Last week of rehabilitation (N = 44)** | **Home – Four weeks after returning home (N = 42)** |
| --- | --- | --- |
|  | **M(SD)** | **M(SD)** |
| FSMC<sup>1</sup> Total | 50.52 (17.69) | 54.71 (17.97) |
| FSMC Motor | 28.02 (9.51) | 30.17 (8.77) |
| FSMC Cognitive | 22.50 (10.10) | 24.55 (10.54) |
| Average Heart Rate (beats per minute) | 75.68 (7.87) | 75.15 (7.65) |
| Steps per Day | 9076.61 (4269.60) | 6826.78 (4329.66) |
| Sleep Duration (minutes) | 444.65 (84.07) | 439.49 (84.43) |
| Time Awake in Bed (minutes) | 29.73 (17.57) | 30.45 (12.65) |
| Intraday Variability | 1.30 (0.22) | 1.29 (0.31) |
| Interday Stability | 0.47 (0.10) | 0.39 (0.10) |
| M10<sup>2</sup> | 5624.62 (8096.23) | 3658.50 (3632.39) |
| L5<sup>3</sup> | 86.88 (254.46) | 124.48 (322.86) |
| Relative Amplitude | 0.95 (0.11) | 0.91 (0.12) |

Note: Mean and standard deviations are reported. Intraday variability and interday stability were calculated using step counts according to the definitions by Taphoorn et al. (1993). <sup>1</sup>FSMC: Fatigue Scale for Motor and Cognitive function, <sup>2</sup>M10: highest activity over ten hours, <sup>3</sup>L5: lowest activity over five hours.

*Objective 1)* Table 3 details the standardized regression coefficients between the derived digital biomarkers and fatigue, which were weak and largely non-significant. Only IV was associated with fatigue significantly (b[95CI] = -0.21[-0.41, -0.02]).

**Table 3.**
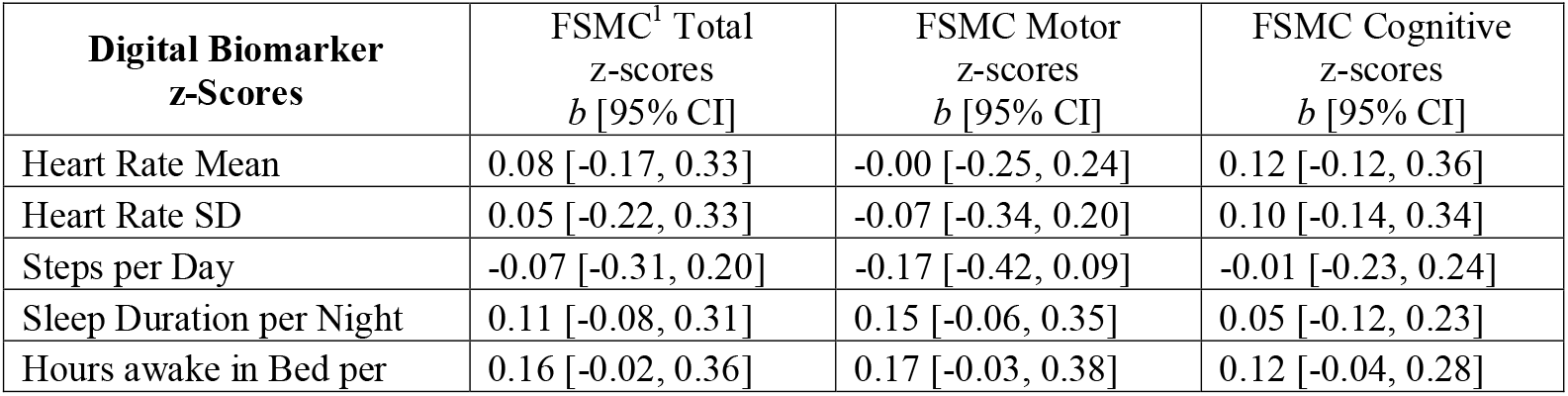
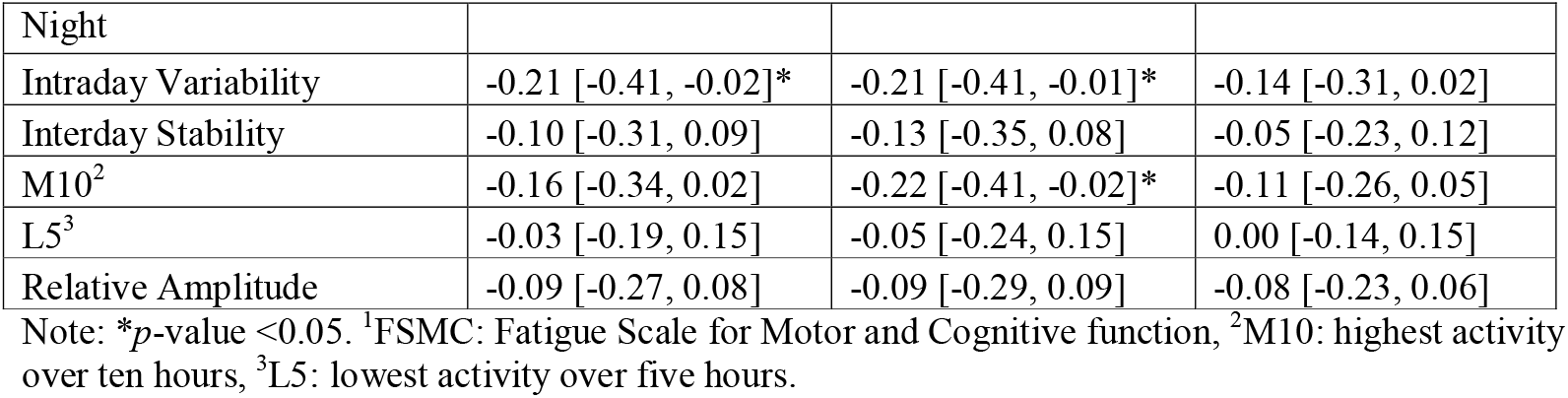
Standardized regression coefficients using FSMC scores as outcome, controlling for age, sex, EDSS status and treatment phase given a random intercept per participant.

### Intraindividual fatigue changes and digital biomarker responsiveness

Average FSMC scores reported in the last week of treatment were 50.5 (SD = 17.7) and 54.7 (SD = 17.9) four weeks after returning home, as illustrated by Fig 1. Sub-scale scores are depicted in S1 Fig in the appendix. The 43 participants who completed FSMC scores at the end of rehabilitation and at the last home follow-up reported an average total fatigue score increase of 4.6 points (SD = 16.6). This rise consisted on average of an increase of 2.0 (SD = 7.8) points in cognitive fatigue and 2.6 (SD = 9.7) points in motor fatigue.

**Fig 1.**
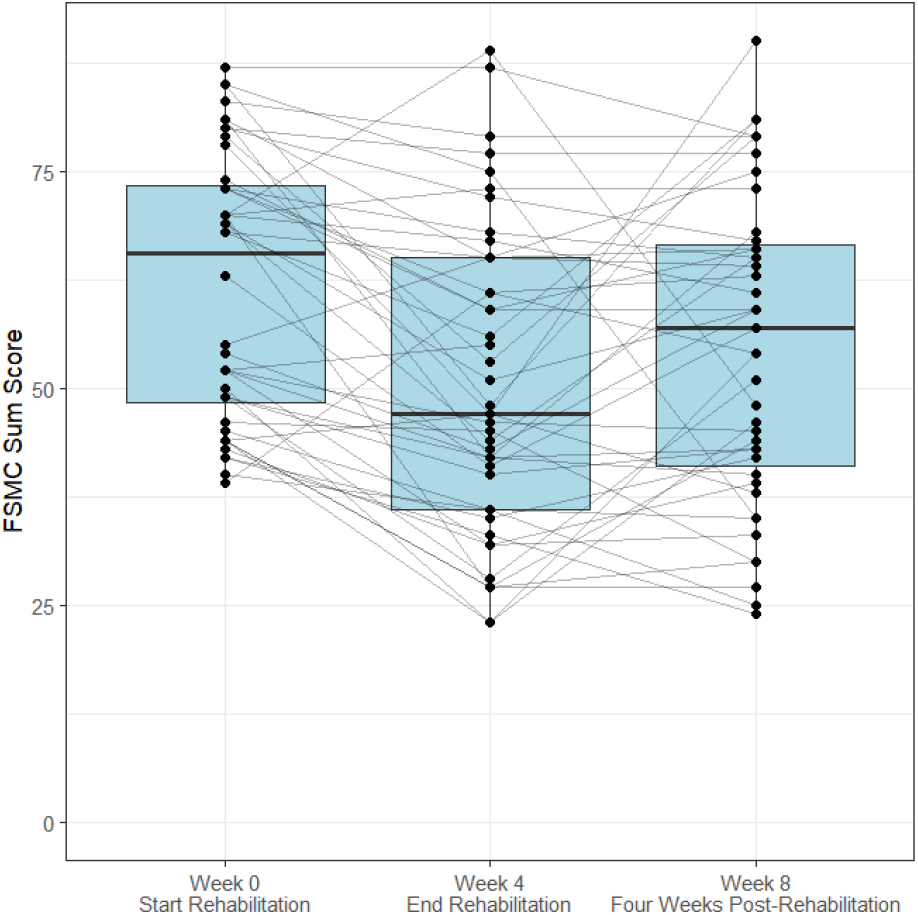
FSMC sum scores at the beginning and end of rehabilitation treatment, as well as four weeks after returning home. FSMC: Fatigue Scale for Motor and Cognitive function.

These changes from end of treatment to four weeks after returning home strongly varied between individuals (n_increase_=23, n_no change_=6, n_decline_=14). In absolute terms, participants experienced on average a shift in total FSMC score of 11.7 points (SD = 12.5), breaking down to 5.4 (SD = 6.0) in cognitive fatigue and 7.4 (SD = 6.7) in motor fatigue.

*Objective 2)* The digital biomarker, which corresponded most closely to changes in fatigue, was daily steps (b = -0.32[-0.68, 0.05]), meaning that increases in fatigue scores by one standard deviation (16.7 points) after returning home were associated with a reduction of 1173.6 in daily steps. Associations of all digital biomarker change scores are reported in Table 4.

**Table 4.** Standardized regression coefficients of changes in digital biomarkers predicting changes FSMC scores, controlling for age, sex, and EDSS status.

| Change in Digital Biomarkers z-Scores | Change in FSMC<sup>1</sup> Total z-score<br>$b$ [95% CI] | Change in FSMC Motor z-score<br>$b$ [95% CI] | FSMC Cognitive Change Score<br>$b$ [95% CI] |
| --- | --- | --- | --- |
| Heart Rate Mean | 0.21 [-0.37, 0.79] | 0.23 [-0.43, 0.88] | 0.16 [-0.31, 0.64] |
| Heart Rate SD | 0.03 [-0.37, 0.43] | 0.00 [-0.44, 0.45] | 0.05 [-0.28, 0.38] |
| Steps | -0.32 [-0.68, 0.05] | -0.36 [-0.77, 0.05] | -0.23 [-0.53, 0.07] |
| Sleep Duration | 0.14 [-0.12, 0.4] | 0.23 [-0.06, 0.51] | 0.05 [-0.17, 0.26] |
| Time Awake in Bed | 0.16 [-0.08, 0.41] | 0.18 [-0.09, 0.46] | 0.12 [-0.09, 0.32] |
| Intraday Variability | -0.11 [-0.39, 0.16] | -0.15 [-0.46, 0.16] | -0.07 [-0.29, 0.16] |
| Interday Stability | 0.01 [-0.25, 0.28] | 0.01 [-0.29, 0.3] | 0.02 [-0.19, 0.23] |
| M10 | -0.19 [-0.42, 0.04] | -0.2 [-0.46, 0.06] | -0.15 [-0.34, 0.04] |
| L5 | -0.08 [-0.28, 0.13] | -0.13 [-0.37, 0.1] | -0.01 [-0.18, 0.16] |
| Relative Amplitude | -0.06 [-0.28, 0.16] | -0.03 [-0.28, 0.22] | -0.08 [-0.26, 0.1] |

Note: <sup>1</sup>FSMC: Fatigue Scale for Motor and Cognitive function, <sup>2</sup>M10: highest activity over ten hours,
<sup>3</sup>L5: lowest activity over five hours.

We further compared the strength of associations we found for within-person change scores in Table 4 to the strength of overall associations we saw in Table 3. Regression coefficients from the change model (i.e., examining differences between time points) were on average 23.3% greater than the coefficients from the cross-sectional model.”

### Cognitive and motor fatigue subscales

*Objective 3)* Overall associations reported in Table 3 were on average 60.1% stronger for the motor fatigue subscale (b = 0.13) compared to the cognitive subscale (b = 0.08). Similarly, associations of digital biomarker changes with changes in fatigue were on average 62.9% stronger for the motor fatigue subscale (b = 0.15) compared to the cognitive subscale (b = 0.09).

Congruent with these trends, the largest effect size of -0.36 [-0.77, 0.05] was observed between changes in steps and changes in the motor fatigue subscale.

## Discussion

This study investigated the utility of wearable devices in fatigue monitoring with a further focus on responsiveness to change and differences between cognitive and motor fatigue. While the observed associations of digital biomarkers and patient-reported fatigue were weak overall, we found some evidence that digital biomarkers were more responsive to fatigue score changes and better corresponded to motor than cognitive fatigue. We speculate that the overall weakness of associations is primarily due to two main constraints. Firstly, objectively observable metrics such as heart rate and step counts are confounded by many biological and behavioral factors (24), which limit how well digital biomarkers can explain subjectively experienced fatigue. We further suspect that the Fitbit Inspire HR measurement accuracy and data aggregation are limiting factors, as these consumer-grade devices are primarily designed for affordable lifestyle tracking instead of clinical-grade physiology measurements.

Despite much interest in digital biomarker monitoring for fatigue management, detecting reliable and accurate digital biomarkers remains a challenge (25,26). Although our analysis has not been able to produce robust evidence, the unique BarKA-MS study setting, including marked changes in fatigue scores over time still enabled us to add to the evidence base. Previous studies conducted purely in free living conditions observed changes at a much slower pace, such as Abdelhak and colleagues (2024), who reported on patient-reported outcomes in progressive MS and found an average increase of 4.9 points on the FSMC scale over 1.6 years (27). Skorve and colleagues (2020) investigated cognitive function changes over two years in recently diagnosed PwMS and observed an FSMC increase of 3.8 points (28). Therefore, we believe that the fluctuations between inpatient rehabilitation and free-living assessment provide an excellent opportunity for fatigue monitoring research and to examine the responsiveness of digital biomarkers.

We were also able to observe some promising preliminary signals, which we believe are worth investigating in future research. Digital biomarkers were consistently more strongly associated with within-person changes in fatigue than cross-sectional fatigue scores. In our opinion, this suggests that digital biomarkers are more suitable for capturing within-person changes than for cross-sectional analysis such as discriminating between fatigued and non-fatigued people. This finding is promising because the most feasible implementation of wearable devices in clinical care for PwMS would likely be in long-term monitoring and assessment of intra-individual trends. The second consistent pattern that emerged in our fatigue sub-scale analysis was that digital biomarkers corresponded better to motor fatigue than cognitive fatigue. This finding is intuitive but, in many studies, cognitive and motor fatigue are not explicitly differentiated. Consistently with both patterns, we conclude that digital biomarkers had the strongest relationships with within-person changes, thus likely offering the most promising outcome for future digital biomarker studies.

We also found some evidence that in cross-sectional analysis individual circadian rhythm patterns, such as interdaily stability and intradaily variability, may be promising digital biomarkers. During rehabilitative treatment the average intradaily variability (IV) and interdaily stability (IS) values (IV = 1.30, IS = 0.47; Table 2) were close to those seen in people in post-stroke rehabilitation (IV = 1.23, IS = 0.48; (29)) and worse that those found in a nationally-representative US sample (IV = 0.69, IS = 0.58;(30)). This comports well with the lived experience of fatigue for many patients who show a severe lack of energy and activity after overexertion, which would result in a fragmented daily activity rhythm. However, it should be noted that this finding is not consistent with a previous on RARs in MS (IV=0.58, IS=0.67; (31)), which found RARs to be more regular in PwMS than in persons without MS. This inconsistency highlights the need for further research with strong external validity.

### Strengths

Our study’s strengths lie in its high data quality and the study setting, which produced marked changes in fatigue between the rehabilitation and the home phase. Thanks to these, we were able to examine the responsiveness of digital biomarkers in a well-documented and highly compliant study population. A further notable contribution is the inclusion of highly burdened PwMS, as this population has been rather under-researched so far. In our sample, 29.5% required walking aids when walking 100 meters, which is particularly relevant given that consumer-grade activity devices, which are not optimized for such gait styles. Many PwMS already use such a device (32), and precisely understanding their utility and limitations is important for their integration into research and clinical care. If digital biomarkers become reliable within a person, these devices could help PwMS in managing their daily activities and avoid over-exertion and the following post-exertional malaise.

## Limitations

Further research involving larger samples is needed to be confident that findings will generalize. Limitations in the granularity and precision of the data may also have inhibited us from identifying clearer digital biomarker signals. For example, our technical set-up was unable to capture heart rate Variability, which has previously been shown to be a very promising biomarker for fatigue (33). Besides IV and IS, the third commonly reported metric is the relative amplitude (RA), which measures the ratio of the highest (M10) to the lowest daily activity (L5). Because the Fitbit Inspire HR only reports full steps and not more raw activity counts reflective of minor movements, the L5 stat was often close to zero, which resulted in the RA being close to its maximum of 1. This reduced variability also limited its utility in this analysis.

It is also important to consider that many external circumstances change simultaneously when patients return home from in-patient treatment, which may lead to confounding.

### Future directions

Recruiting larger samples and using research-grade devices would likely lead to stronger signals and more reliable findings. Due to the high resource demands of such data collection, we believe that pooling and reanalyzing existing data should also be explored as a supplemental approach. Larger pooled datasets would offer greater generalizability and allow for more advanced machine learning methods to identify more sophisticated biomarkers. Consequently, the data used in this study are made openly available to support such efforts (34).

## Conclusion

This study finds that digital biomarkers, based on consumer-grade activity watches, struggle to capture fatigue in people with high MS symptom burden, even if the data is collected very consistently. In cross-sectional analyses rest-activity rhythms may be among the most promising digital biomarkers. However, overall our findings suggest that digital biomarkers are better suited to reflect longitudinal changes and are more closely linked to motor than to cognitive fatigue. We speculate that future work focusing on these aspects would enjoy the best chances of success in identifying reliable digital biomarkers.

## Data Availability

The data underlying this manuscript is made openly available through a Zenodo repository.

https://doi.org/10.5281/zenodo.17485784

