## Supplemental Figure 1 for "Monitoring fatigue with consumer wearables in multiple sclerosis following rehabilitation: An observational study"

Supplementary Materials

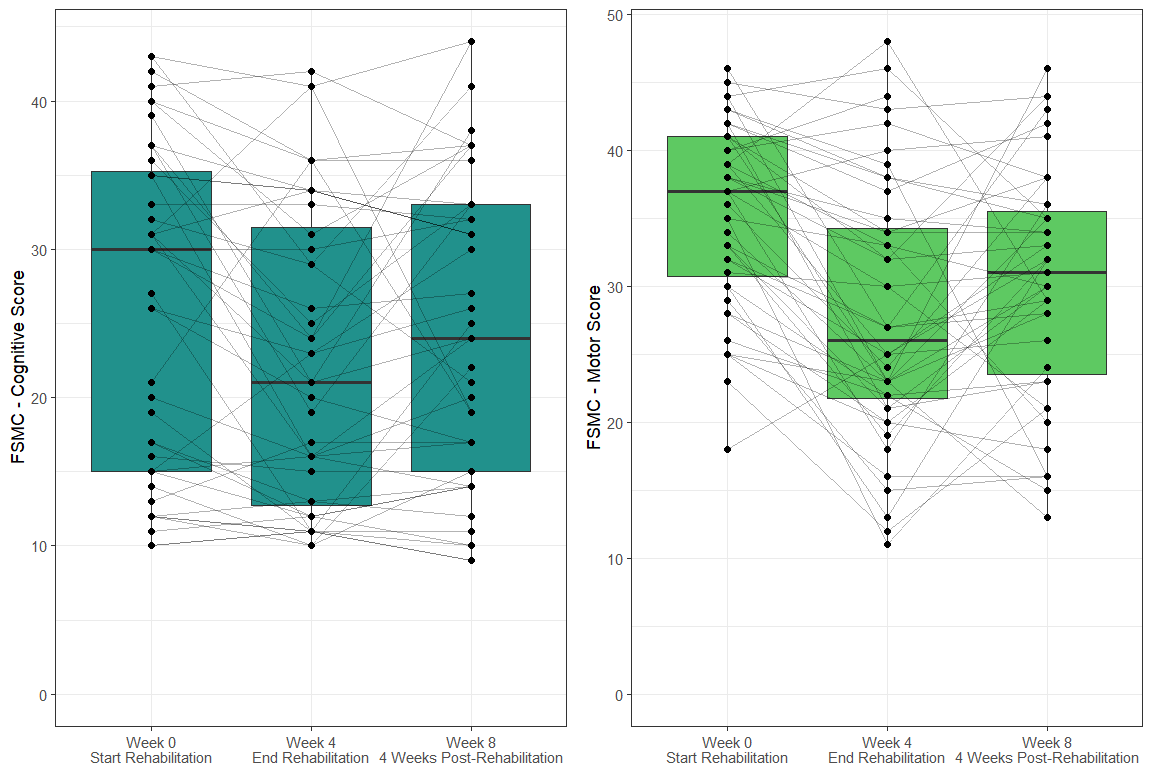

**S1 Fig.** **FSMC sub-scale scores at the start and end of rehabilitation as well as four weeks after discharge.** FSMC: Fatigue Scale for Motor and Cognitive function.
